# Estimating real-world COVID-19 vaccine effectiveness in Israel using aggregated counts

**DOI:** 10.1101/2021.02.05.21251139

**Authors:** Dvir Aran

## Abstract

The vaccination roll-out of the COVID-19 vaccines in Israel has been highly successful. By February 22th, approximately 47% of the population has already been administered at least one dose of the BNT162b2 vaccine. Efforts to estimate the true real-world effectiveness of the vaccine have been hampered by disease dynamics and social-economic discrepancies. Here, using counts of positive and hospitalized cases of vaccinated individuals, we conduct a sensitivity analysis of the vaccine effectiveness. Under an assumption of no effectiveness on the first two weeks after the 1st dose, we observe very low effectiveness on the third week. After the 2nd dose, on weeks 1 and 2 we find 73-85% effectiveness in reducing positive cases, hospitalizations, and severe cases, which increase to 89-97% effectiveness 14 days after the 2nd dose. As more granular data will be available, it will be possible to extract more exact estimates; however, the emerging evidence suggests that the vaccine is highly effective.

## Introduction

Vaccination rollout in Israel of the COVID-19 vaccines started on December 20, 2020. By February 9th, 47% and 32% of the population had already received the 1^st^ dose and 2^nd^ dose, respectively, by the BNT162b2 vaccine developed by BioNTech and Pfizer. The vaccination campaign coincided with the beginning of a “3rd wave” of infections, and by mid-January SARS-CoV2 positive cases and hospitalizations more than doubled. To mitigate this increase in cases, on January 8 a strict lockdown was imposed. However, cases and hospitalizations did not drop as expected and as observed in previous waves. There was some frustration in the public and by government officials, and doubts were raised whether the vaccines are effective.^1^

Estimating real-world effectiveness of vaccinations is complicated compared to a randomized, controlled and double-blinded clinical trial. First, in real-world there is no control group. With the protection from the vaccine cases are eliminated, and the general population incidence does not represent the incidence rate with no vaccinations. Second, in real-world there is no randomization. Israel has seen significant discrepancies between socio-economic and demographics groups in vaccination uptake.^2^ Additionally, COVID-19 disproportionately stroked individuals of lower socio-economic status. Third, the real-world vaccination is not blinded. Behavioral changes of those immunized may affect the number of encounters and chances of infection. In summary, while in the clinical trial the disease dynamics, socio-economic differences and behavioral aspects can all be controlled, in real-world, it is more complicated to accurately tease out those confounding factors.

Here, using publicly available data of COVID-19 dynamics and SARS-CoV2 positive and hospitalizations of those that were vaccinated, we provide estimates of the effectiveness of the vaccine in reducing cases, hospitalizations and severe cases due to COVID-19. All data and code used in this study are available at https://github.com/dviraran/covid_analyses.

## Methods

Daily SARS-CoV2 positive cases and numbers of severe or critical hospitalization were downloaded from the Israeli Ministry of Health (MoH) COVID-19 public database.^3^ Number of positives cases, hospitalizations and severe or critical hospitalizations of vaccinated individuals was provided by the MoH on February 23^th^, 2021. The counts are stratified by ages 60 years and above (60+) and below 60 years (60-), and five groups according to number of days from the vaccination: between day 0 to 13 of the first dose (group 1), between day 14 to 20 of the first dose (group 2), between day 0 to 6 of the 2^nd^ dose (group 3), from day 7-13 of the 2^nd^ dose (group 4), and from the 14+ (group 5).

To calculate vaccine effectiveness (VE), we first estimate the expected number of cases or hospitalizations (**Supplementary Figure 1**). To achieve this, we count the number of the cumulative vaccinated individuals on each day that are eligible in each group. We call this vector V(g), where g is the relevant group as described above. We then calculate the daily incidence rate of cases in the whole population. Naively, this is the number of cases divided by the population size. However, the daily incidence (*d*) is affected by the VE, as there are cases that are eliminated by the vaccination. To overcome this issue, we deconvolve the number of observed cases in the population per day – the number of cases from those vaccinated after the 2^nd^ dose (*S*^1^) and those not vaccinated or before 2^nd^ dose (*S*^2^). Those vaccinated are multiplied by 1 minus the VE, which provides the number of cases if they were not vaccinated. Finally, since incidence rates of the vaccinated cohort are different from the general population, we use a sensitivity parameter *β* to adjust for the incidence rates. Based on all this, the formula for the VE is as below:

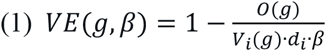

And the daily incidence *d*_*i*_ can be calculated by the sum of 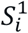 and 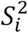:

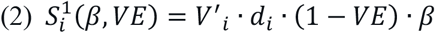

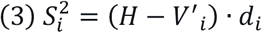

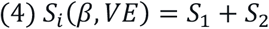

Where *O*(*g*) is the observed number of cases in the relevant group; *S*_*i*_ is the number of cases on day i; *V* ′_*i*_ is the number of vaccinated individuals after the 2^nd^ dose on day i; *d*_*i*_ is the daily incidence; *VE* is the effectiveness of the vaccine; *β* is the sensitivity parameter; and *H* is the population size (1,428,000 for >60 years old, 7,539,000 for <60 years old).

To find the solution of eq. 1 for VE, we find the value of VE by minimizing the following function, which is a combination of eq. 1 and 4:

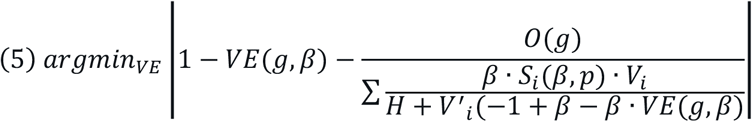

To estimate *β* for each group, we hypothesized that by day 13 of the 1^st^ dose, there should not be an observed effect of the vaccine. We present ±10% of the *β* values.

## Results

Between December 20, 2020 and February 23th, 2021, there were 4,295,685 individuals vaccinated in Israel by the 1^st^ dose of the BNT162b2 vaccine, of them, 1,303,244 over the age of 60 (91% of the whole 60 years and over population). By that date, 2,918,008 have already received their 2^nd^ dose of the vaccine. Of all those vaccinated, 52,014 individuals have tested positive for SARS-CoV2, 3,148 have been hospitalized due to COVID-19 and 2,141 were hospitalized with severe or critical conditions or have died (**Table 1**).

**Table 1.** Number of cases of vaccinated individuals as reported by the Ministry of Health of Israel.

|  | *Positive cases (>60y)* | *Positive cases (<60y)* | *Hospitalizations (>60y)* | *Hospitalizations (<60y)* | *Severe (>60y)* | *Severe (<60y)* |
| --- | --- | --- | --- | --- | --- | --- |
| *1<sup>st</sup> dose, day 0-13* | 7,743 | 24,104 | 1,262 | 426 | 936 | 215 |
| *1<sup>st</sup> dose, day 14-20* | 5,532 | 8,447 | 826 | 119 | 595 | 52 |
| *2<sup>nd</sup> dose, day 0-6* | 1,254 | 1,573 | 171 | 27 | 132 | 11 |
| *2<sup>nd</sup> dose, day 7-13* | 1,276 | 1,009 | 196 | 20 | 137 | 11 |
| *2<sup>nd</sup> dose, day 14+* | 629 | 447 | 88 | 13 | 46 | 6 |

Based on daily numbers of vaccinations and rates of general incidence we estimated expected numbers of SARS-CoV-2 positive cases, COVID-19 hospitalizations and severe cases. We correct our estimation to two confounding factors. First, the incidence rates of those that were vaccinated early are not similar to the general population, as previous analyses have shown that older populations have lower incidence and lower socio-economic groups have higher incidence.^4^ Therefore, we perform a sensitivity analysis by adjusting incidence rates using different levels of *β* values. Second, the general incidence rate is affected by the vaccinations, as individuals that have been vaccinated would have been infected without the vaccine. Therefore, we derived a formula that estimates the daily incidence rate by adding those eliminated cases as a function of the VE (**Methods**).

For positive cases, which are a combination of symptomatic and asymptomatic individuals, in ages 60 years and above, we find the empirical *β* to be 0.78, which implies that the vaccinated population are expected to have 78% the cases of the general 60+ population (**Figure 1A**). Strikingly, the analysis suggests no effect at all by day 20 of the 1^st^ dose. However, on the 4^th^ and 5^th^ week, which are also weeks 1 and 2 after the 2^nd^ dose, we observe a reduction of 73%, which increase to 96% reduction from 14+ days and above after the 2^nd^ dose. For individuals aged below 60 years, the empirical beta values are 0.81. Here we see reduction 77% after the 2^nd^ dose, 81% reduction in week 5, and 94% reduction from 14+ days and above after the 2^nd^ dose (**Figure 1B**).

**Figure 1.**
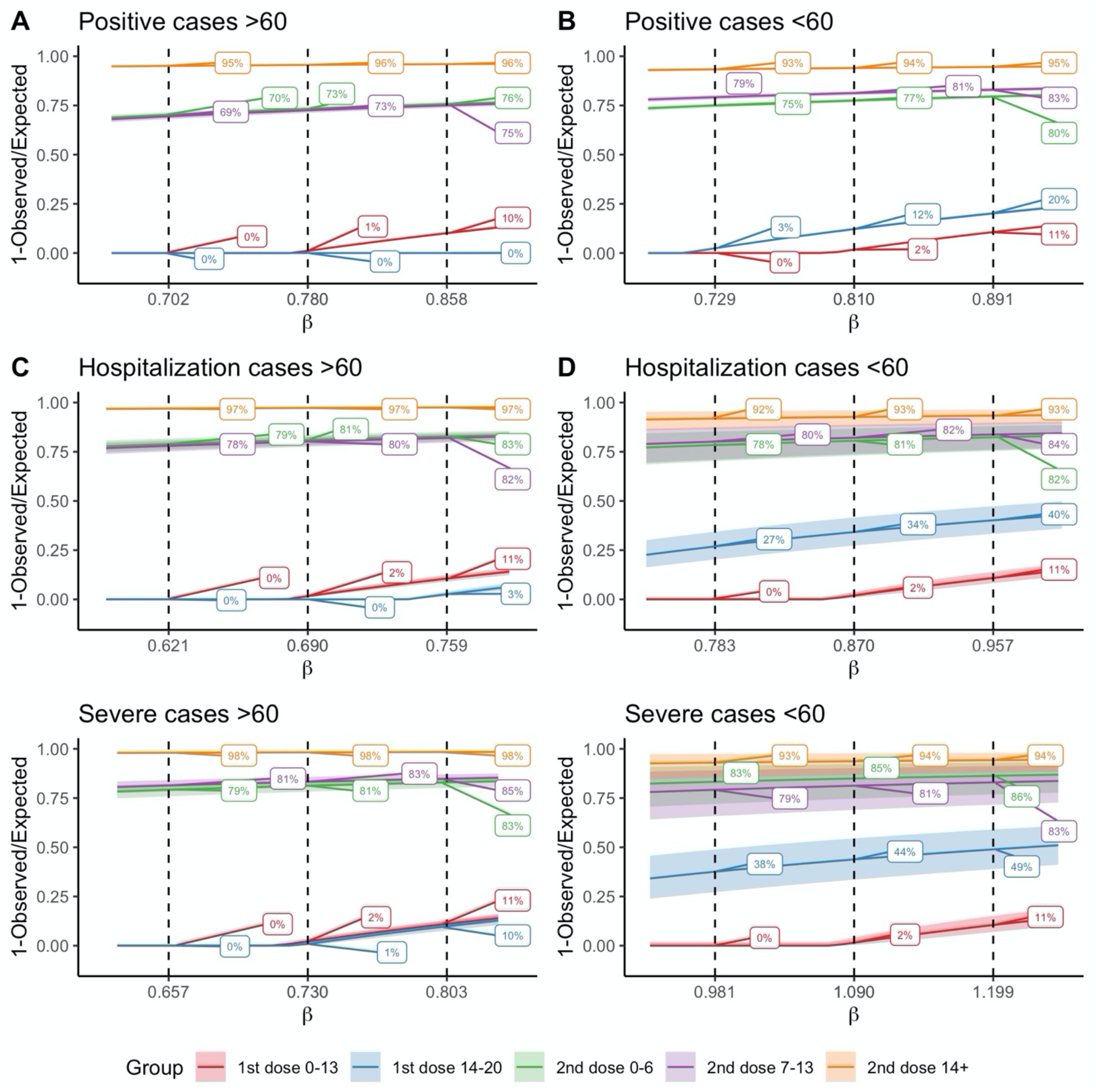
Effectiveness rate estimations of the vaccination by different levels of beta values. Each plot shows the estimated effectiveness (y-axis) as a function of β. Dashed lines are for empirical β value, and +-10% from the β value. 95% Confidence intervals are in shade.

Similarly, we perform the analysis for hospitalizations and severe cases, stratified by the age groups. For hospitalizations in 60+ our analysis suggests beta values of 0.69. Similarly, we do not observe any effect by day 20. After the 2^nd^ dose we find 79-81% reduction in hospitalizations, which increase to 97% reduction two weeks after the 2^nd^ dose (**Figure 1C**). We observe similar effectiveness in the 60-group, however with large confidence intervals, as numbers of hospitalizations are low. Interestingly, in this group we do observe low but not negligible VE of 34% in the third week (**Figure 1D**).

Finally, in severe case we estimate the *β* values to be between 0.73. Up to day 20 we observe no reduction in severe cases, but after the 2^nd^ dose we observe a reduction of 81-83% in the first two weeks and 98% after day 14 (**Figure 1E**). Similar to hospitalizations, in those below age 60, the number of severe cases is too low for a reliable estimation, but the trend is similar to the 60+ group. Again, we observe non negligible VE of 44% on the third week.

It is important to note, that while positive cases may already have been counted, hospitalizations and severe cases may deteriorate later and the number of cases is expected to increase, which in turn will reduce the estimation of the VE. To test whether this censoring affects the estimations we performed a censoring analysis on the vaccinated individuals in the group of 14+. The analysis shows that even when taking into account only those with 20+ days after the 2^nd^ dose for calculating the expected number of cases, the VE for 60+ is still 91% for positive cases, 95% for hospitalizations and 97% for severe disease (**Supplementary Figure 2**).

## Discussion

The randomized clinical trial (RCT) of BNT162b2 has suggested efficacy of 95% a week after the 2^nd^ dose and unclear efficacy earlier.^5^ It also suggested differences between the older and younger population, but with large standard errors due to relatively small sample sizes. In addition, the clinical trial was performed on a relatively small population; in contrast, by February 23^th^, in Israel alone 195-fold more individuals have been vaccinated compared to the trial. Therefore, real-world data VE is of high interest and important for decision-makers and mobilizing individuals to get the vaccine. Our real-world estimates are very similar to the RCT, however with a few differences. First, we do not observe any VE before the second dose. Second, full VE is only observed 14 days after the 2^nd^ dose, and not 7 days as suggested by the RCT.

Our method adjusts for two main issues in comparing cases in vaccinated individuals and the general public. The sensitivity analysis provides VE estimates that corrects for demographic, socio-economic and additional behavioral aspects. Our deconvolution approach for correcting incidence rates allows to estimate VE in situations where the vaccine is already impactful on cases. We report here that two weeks after the 2^nd^ dose the vaccine is over 90% effective in reducing positive cases in individuals older than 60 years, and over 95% effective in reducing hospitalizations and in preventing severe cases.

Our analysis suggest that the vaccine does not provide substantial protection in days 14-20 after the 1^st^ dose, as we only observe substantive effectiveness in days 0-6 of the 2^nd^ dose, which is administered in Israel on the 21st day after the 1^st^ dose of the vaccine. We cannot differentiate here between the possibility that the 1^st^ dose is effective but only after three weeks, or that the vaccine is only protective following the 2^nd^ dose of the vaccine. However, there is some preliminary evidence to support that the single dose is effective after three weeks.^6^

In Israel, individuals may get tested for SARS-CoV-2 for any reason, not just due to symptoms. Thus, the positive cases come from both symptomatic and asymptomatic individuals. This is different from the clinical trial, where only symptomatic individuals with suspected COVID-19 where tested. It might explain some of the difference in effectiveness we observe in Israel regarding positive cases. The lower, VE we observe for hospitalization and severe cases may be explained by the inclusion of diverse population, including older population (the clinical trial was limited to age 85) and more comorbidities.

It is important to note that our estimates of VE in reducing the disease should not be confused with effectiveness in reducing transmission. As noted, we cannot exclude the possibility that vaccinated individuals may still get infected by SARS-CoV-2 and stay asymptomatic or with mild symptoms and will therefore not get tested. However, other studies have shown reduction in Ct values of the PCR test due to the vaccination, suggesting lower viral load, and in turn reduced transmission.^7^

Our analysis suffers from limitations. First, all analyses are performed on aggregated counts, which limits the possibilities to make individual-level inferences. Second, hospitalizations and severe cases may accumulate with time, as some of the patients will deteriorate later on. The number of new vaccinated individuals have been relatively low in the fifth week therefore this issue should not have a major effect on our estimates up to the fifth week. Third, in Israel there is an incentive to get tested if you are required to be in isolation due to contact with an infected individual, and as noted above some asymptomatic individuals are thus identified. However, this incentive is reduced for people who are 7 days after the 2^nd^ dose, as the Israeli MOH regulations now exempts these people from mandated isolation. Thus, there is a difference in testing rates of asymptomatic individuals between groups. It is reassuring that there are relatively similar levels of effectiveness of those 7 days after the 2^nd^ dose to those before those 7 days, suggesting that this testing incentive has only a marginal effect.

Other attempts to identify the impact of the vaccination campaign in Israel are underway. Chodik et al. compared cases in vaccinated individuals on days 13-24 after the 1^st^ dose with vaccinated individuals in days 0-12.^8^ Rossman et al. used a natural experiment approach to compare early and late vaccinated cities and differences in the prioritization for the vaccine between age groups.^4^ Our contribution here is the use of the general population as a control group to assess the effectiveness rather than vaccine impact, and introducing methodological advancements to provide improved real-world estimations.

In conclusion, this study provides estimate the effectiveness of the BNT162b2 vaccine on a population level compared to the general population. Our analysis provides strong reassurance that the vaccine is highly effective. With more data that will be shared with the public we believe that more accurate estimation can be calculated.

## Data Availability

All data is public and can be downloaded from https://data.gov.il/dataset/covid-19 or Ministry of Health press releases. Data and code used in the analyses was deposited in https://github.com/dviraran/covid_analyses. In addition, a shiny app is available, which will be updated as more data is available by the Ministry of Health - https://dviraran.shinyapps.io/VaccineEffectIsrael/.

https://data.gov.il/dataset/covid-19

https://dviraran.shinyapps.io/VaccineEffectIsrael/

## Data and code availability

All data is public and can be downloaded from https://data.gov.il/dataset/covid-19 or Ministry of Health press releases. Data and code used in the analyses was deposited in https://github.com/dviraran/covid_analyses. In addition, we provide an interactive shiny app, which will be updated as more data is available by the Ministry of Health -https://dviraran.shinyapps.io/VaccineEffectIsrael/.

## Acknowledgments

DA is supported by the Azrieli Faculty Fellowship and is a Deloro Fellow. We thank Michael Geller for help in developing the incidence rate formula. We also thank Hagai Rossman for providing accurate data and Uri Shalit for his valuable comments.

## Supplementary Figures

**Supplementary Figure 1.**
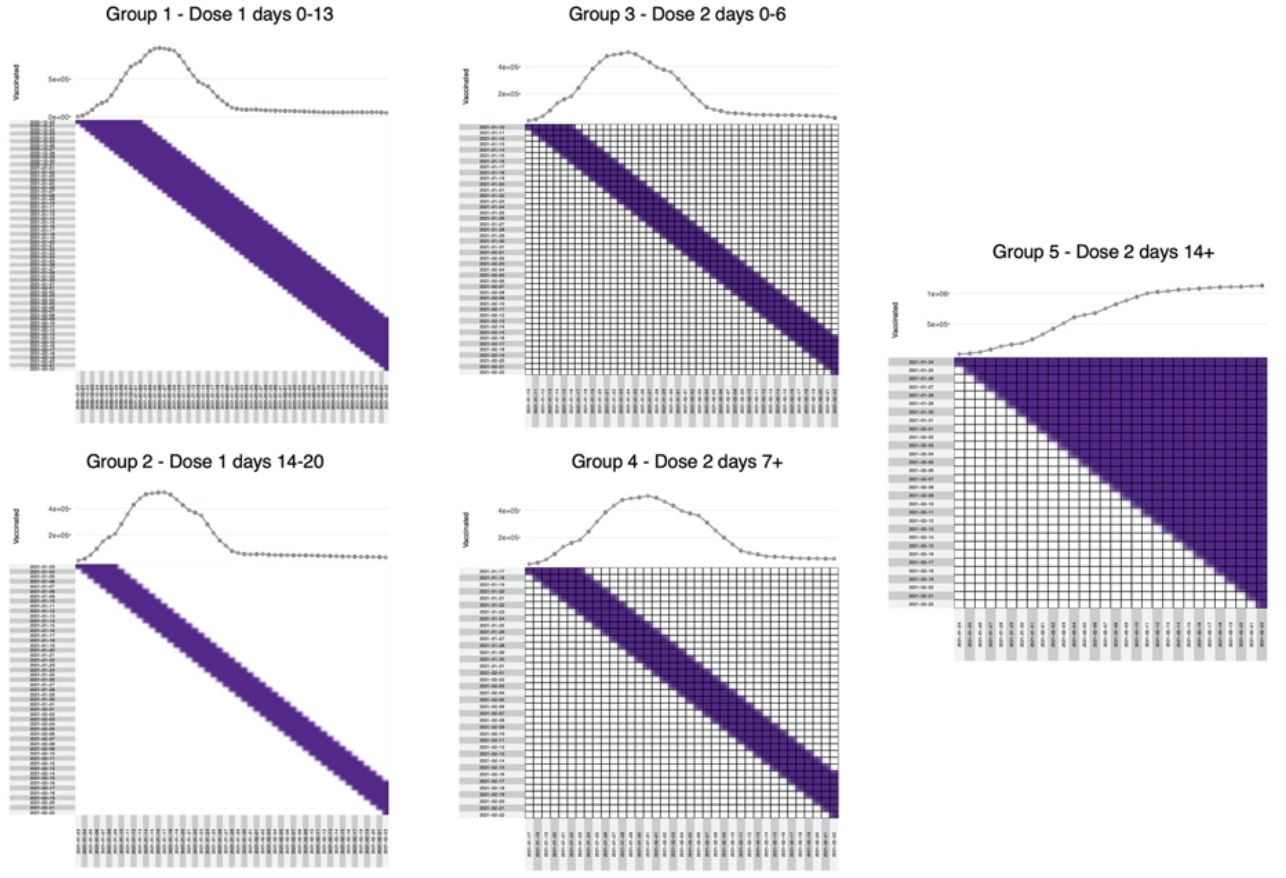
Visualization of expected case counts model. Days counted from vaccination. Columns are days of vaccination; Rows are days of possible infection. A cell is blue if the vaccinated individual is counted in the relevant group. The distribution on the top is the sum of each column (the number of vaccinated individuals that are counted for that date). Note that in group 5 numbers are steadily increasing, and most cases are recent, thus, hospitalizations and severe cases are expected to increase.

**Supplementary Figure 2.**
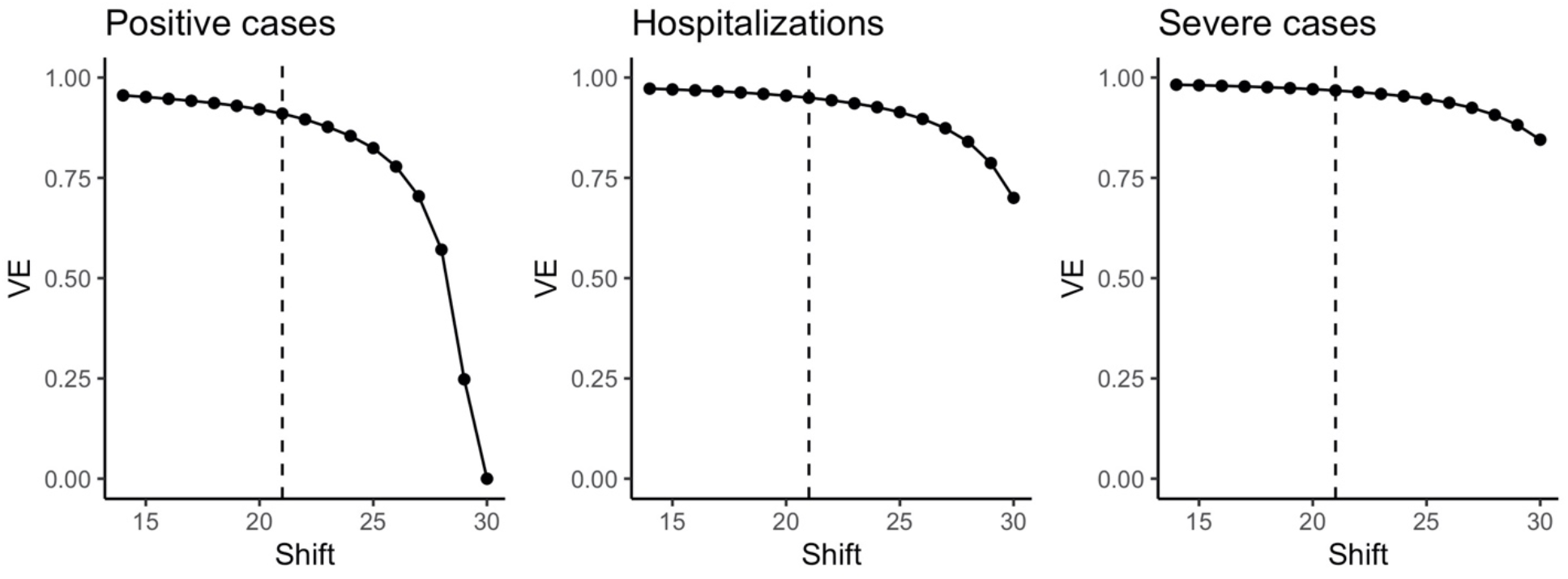
Censoring analysis for 14+ days after 2^nd^ dose group. Vaccine effectiveness is calculated using observed counts of 14+ days after 2^nd^ dose, however, expected counts are calculated using those vaccinated only after number of days in the x-axis. This analysis allows to estimate VE only for those with sufficient time to deteriorate to severe disease.

